# Physical Activity, Sleep, and Quality of Life in Pulmonary Arterial Hypertension: Novel Insights from Wearable Devices

**DOI:** 10.1101/2024.09.30.24314671

**Authors:** Andrew M. Hughes, Alisha Lindsey, Jeffrey Annis, Kelly Burke, Hiral Master, Luke G. Silverman-Lloyd, Jonah D. Garry, Michael J. Blaha, Erika S. Berman Rosenzweig, Robert P. Frantz, Paul M. Hassoun, Evelyn M. Horn, Jane A. Leopold, Franz P. Rischard, Brett Larive, Nicholas S. Hill, Serpil C. Erzurum, Gerald J. Beck, Anna R. Hemnes, Evan L. Brittain

## Abstract

**Background:** Reduced functional capacity and poor sleep quality are common in pulmonary arterial hypertension (PAH). We aimed to determine whether Fitbit-derived activity and sleep trends provide clinically meaningful information in patients with PAH.

**Methods:** Our prospective observational study recruited patients with PAH from across the United States. Participants with PAH wore a Fitbit device for 12-weeks at baseline and a subgroup with one-year follow-up. A matched control cohort was generated from the *All of Us* Research Program and we evaluated changes in patients with PAH compared to matched controls.

**Results:** Among 109 patients with baseline monitoring, average daily steps correlated with 6MWD (r=0.66, p < 0.001) and percent rapid eye movement (REM) sleep (r=0.26, p=0.016). In 44 PAH participants who completed baseline and one-year monitoring, there was a group-time interaction for percent light sleep (p=0.024) and percent REM sleep (p=0.034), which demonstrated that sleep quality worsened in patients with PAH over one year compared to matched controls. Average daily steps in patients with PAH decreased from 5200 [IQR 3212 – 7458] at baseline to 4651 [IQR 2912 – 6827] at one year (p=0.008). Compared to matched controls, activity levels were significantly lower in patients with PAH at both time points.

**Conclusions:** Our study demonstrated the potential clinical value of wearable devices by showing that activity and sleep quality are reduced in PAH compared to matched controls and these measures decline over time. Future studies should investigate if monitoring these health behaviors detects early functional decline and whether targeted interventions may improve outcomes.

**Prior Abstract Publication/Presentation:** Some of the data presented in this manuscript have been included in the following abstracts:

1. Hughes A, Annis J, Hemnes A, Lindsey A, Burke K, Brittain E, and the PVDOMICS Study Group. Sleep Patterns and Quality of Life in Pulmonary Arterial Hypertension. Presented at the American Heart Association (AHA) Scientific Sessions, November 2023, Philadelphia, PA.
2. Hughes A, Annis J, Hemnes A, Lindsey A, Burke K, Horn EM, Brittain E, and the PVDOMICS Study Group. The Role of Long-Term Daily Activity Monitoring for the Assessment of Functional Capacity in Pulmonary Arterial Hypertension. Presented at the American Thoracic Society International Conference, May 2023, Washington D.C.
3. Hughes A, Annis J, Hemnes A, Lindsey A, Burke K, Horn EM, Brittain E, and the PVDOMICS Study Group. Long-Term Daily Activity is Associated with Exercise Capacity and Quality of Life in Pulmonary Arterial Hypertension. Presented at the American Heart Association (AHA) Scientific Sessions, November 2022, Chicago, IL.

## Introduction

Pulmonary arterial hypertension (PAH) is characterized clinically by dyspnea, exercise intolerance, and reduced quality of life (QOL)^1,2^. Reduced functional capacity is a hallmark of PAH. The 6 minute walk test (6MWT) is the most common metric for assessing functional capacity but it has important limitations as a prognostic tool and trial endpoint^3^. Wearable devices, such as activity trackers and smartwatches, are popular among patients and report real-world physical activity and sleep behaviors. Unlike the 6MWT, daily activity integrates physical, environmental, and behavioral inputs of capacity. Remote activity monitoring captures activity over weeks to months, which mitigates the temporary influences of clinic-based assessments. Changes in wearables-derived activity levels correlate with changes in 6-minute walk distance (6MWD) and health-related QOL in short-term studies of patients with PAH^4,5^.

Poor sleep quality is common and associated with mood disorders and impaired QOL in patients with pulmonary hypertension^6,7^, though prior studies have largely assessed sleep quality through self-reported qualitative surveys. Studies with objective, longitudinal sleep metrics are lacking in PAH. Wearable devices represent an emerging low-cost, user-friendly tool to assess sleep duration and quality by providing information on the amount of time that individuals spend in different sleep stages^8,9^.

We aimed to study the objective, free-living data from commercially available wearables to understand the relationship between sleep, physical activity, and QOL in patients with PAH over the course of one year. This study was designed to reflect real-world behavior trends rather than investigate new perspectives on exercise physiology in PAH. Our study is the first in the PAH population to describe longitudinal trajectories of sleep quality with sleep-stage data, provide novel insights regarding longitudinal changes in physical activity, and directly correlate sleep and activity in PAH and matched controls.

## Methods

### Study Design

This is a prospective observational study of patients with PAH between October 2019 and December 2023. This study was approved by the Vanderbilt University Medical Center Institutional Review Board. Matched controls were identified from the All of Us Research Program (AoURP)^10^. Only authors who completed the AoURP Responsible Conduct of Research Training handled the data on the secured cloud platform – Researcher Workbench, according to AoURP policy.

### Study Population

The study enrolled adults in the United States with PAH confirmed by hemodynamics and expert clinical diagnosis. We included participants from several recruitment sources. Forty-three participants were recruited from the NHLBI-funded Longitudinal Pulmonary Vascular Disease Phenomics Program (L-PVDOMICS), a sub-study of the Pulmonary Vascular Disease Phenomics Program^11^. An additional 22 participants from a previously published trial^12^ with a similar 12 weeks of observational monitoring were included in the baseline correlation analyses, but this trial design did not have follow-up. Additional enrollment sources included patients treated in our clinic and patients who attended the Pulmonary Hypertension Association Research Room, the latter of which captures patients who are treated throughout the United States. Seventeen participants were enrolled from the research room and medical records for these participants were reviewed by study investigators to confirm PAH diagnosis. Our ability to enroll participants from several sources led to a diverse, “multicenter” cohort of patients with PAH. Participants were excluded if they were hospitalized within the last 3 months, pregnant, or had orthopedic limitations that precluded 6MWD testing.

The matched control cohort was derived from All of Us Research Program (AoURP) participants in the “Bring Your Own Device” Program who linked their personally-owned Fitbit devices with their electronic health record. Mahalanobis distance matching created the matched control cohort in a 3:1 fashion based on age, sex, body mass index (BMI), and Fitbit monitoring dates. Controls were matched with the 44 patients with PAH who completed both the baseline and one-year follow-up monitoring period. The study used the Controlled Tier Dataset (version 7). The AoURP and Fitbit data have been described previously^13^.

### Baseline Assessment

The baseline assessment included 12 weeks of activity and sleep monitoring with a Fitbit device, 6MWD, World Health Organization (WHO) functional class assessment, PAH medications, and QOL surveys. Participants wore the Fitbit device 24 hours per day for 12 weeks except for charging the device or while bathing. The 12-week study duration was selected to optimize compliance and avoid excessive participant burden, while minimizing any potential Hawthorne effect that may occur during the initial stages of monitoring. 6MWD and functional class assessment were completed as part of the L-PVDOMICS enrollment protocol. For participants enrolled outside of L-PVDOMICS, the 6MWD and functional class were included in analyses if they were completed during a clinic visit within 6 months of the baseline activity monitoring. In a minority of patients enrolled outside L-PVDOMICS, the WHO functional class was not stated in the available medical records, so these individuals were characterized as unspecified. QOL was assessed through the Minnesota Living with Heart Failure (MLHF) questionnaire^14,15^. The MLHF questionnaire is a 21-item survey scored on a Likert scale (0-5), which calculates a total score (21 questions, range 0-105), a physical score (8 questions, range 0-40), and an emotional score (5 questions, range 0-25). Since the survey includes eight questions that only contribute to the total score, it is possible to have worsened QOL by the Total MLHF score without significant changes in the MLHF physical or MLHF emotional score. Higher MLHF scores indicate greater impairment.

### Follow-Up Assessment

At one year follow-up, participants wore the Fitbit device for an additional 12 weeks and completed the online surveys to capture MLHF scores and PAH medications. 6MWD and functional class were not re-assessed at follow-up.

### Devices and Activity/Sleep Data Capture

Fitbit devices collected activity and sleep data in all participants. Multiple models were used throughout the study including: Fitbit Charge 3, Fitbit Charge 5, and Fitbit Versa. The same Fitbit model was used for the entire duration of a 12-week monitoring period. The Fitbit device was mailed to the participant and a coordinator assisted the participant with set-up over the phone.

The Fitbit device collects several activity metrics including steps per day and total minutes of activity intensities per day (sedentary, lightly active, fairly active, and very active), which are based on calculated metabolic equivalents through proprietary algorithms. A valid day of physical activity monitoring required at least 100 steps and 10 hours of wear time. Fitbit activity metrics were measured daily for approximately 12 weeks and averaged over the entire monitoring period. Sleep variables included total minutes asleep and percent of time in each sleep stage, which was calculated as minutes in each stage divided by total sleep duration. Fitbit devices use accelerometers and photoplethysmography to capture the sleep variables via proprietary algorithms. Participants with fewer than 15 total days of sleep tracking in a monitoring period were removed from analyses.

Activity and sleep data were collected in real-time. Automated text message reminders were sent to participants if there was no data transmitted to the data collection program (Fitabase) after 24 hours. Coordinators contacted participants via telephone if more than 72 hours elapsed since their last data transmission. Participants were unblinded to their activity and sleep levels.

### Outcome Measures

We investigated associations between Fitbit-derived activity and sleep measures, 6MWD, and QOL in patients with PAH and studied how these metrics changed over one year. We directly compared activity and sleep levels in patients with PAH to matched controls at baseline and one-year follow-up.

### Statistical Analyses

Continuous variables are expressed as median with interquartile range (IQR) and categorical variables as counts with proportions. The Wilcoxon signed-rank test compared continuous variables at baseline and one-year follow-up in the PAH cohort. The Wilcoxon rank-sum test compared continuous variables between PAH patients and matched controls. Chi-Square tests compared categorical variables. Associations between activity and sleep metrics were analyzed with the Spearman’s rank correlation test. Generalized least squares models, adjusted for age, sex, and BMI, with linear interaction terms evaluated changes in sleep variables from baseline to year 1 in patients with PAH compared to matched controls. All analyses were conducted in the R programming language (version 4.3). Matched case control analyses specifically utilized the Researcher Workbench within the same R environment. Generalized least squares models were fit using the rms R package. Mahalanobis distance matching was performed in the MatchIt R package.

## Results

### Patient Characteristics

109 participants completed the 12-week baseline monitoring period. The cohort was median age 52.7 years (IQR: 40.9 – 60.6), female predominant (85% female), and majority WHO functional class II. The most common PAH etiology was idiopathic (61%) with participants taking an average of 1.5 PAH medications at the time of enrollment **(Table 1)**.

**Table 1:**
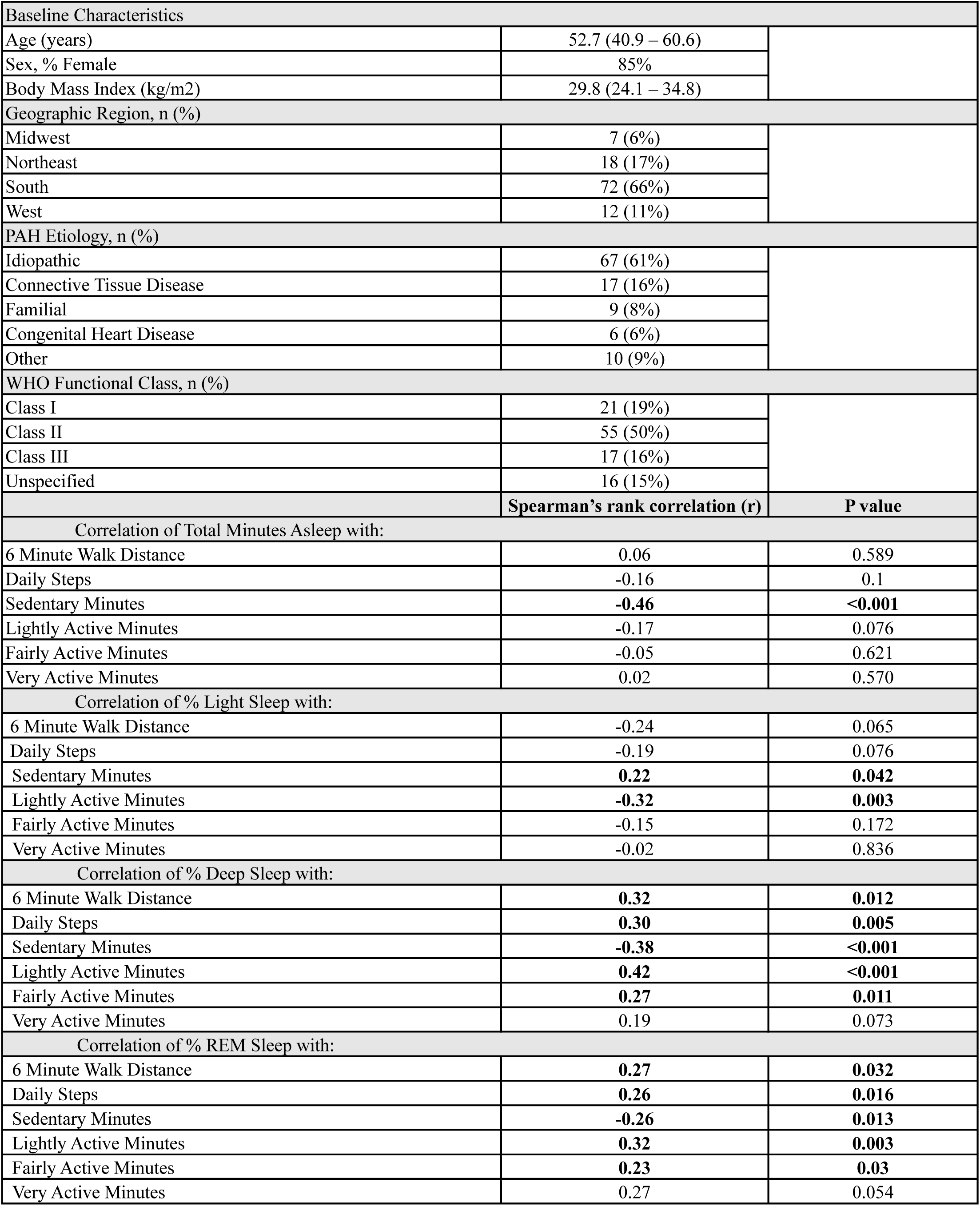
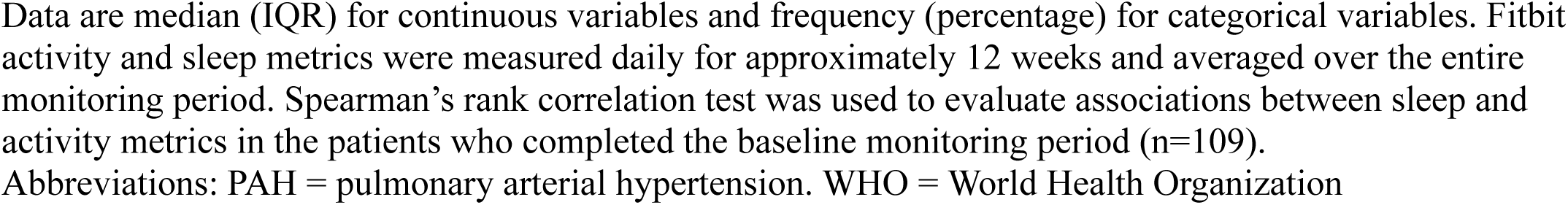
Correlations between Sleep and Activity Measures in PAH (n= 109)

| Baseline Characteristics |  |  |
| --- | --- | --- |
| Age (years) | 52.7 (40.9 – 60.6) |  |
| Sex, % Female | 85% |  |
| Body Mass Index (kg/m2) | 29.8 (24.1 – 34.8) |  |
| Geographic Region, n (%) |  |  |
| Midwest | 7 (6%) |  |
| Northeast | 18 (17%) |  |
| South | 72 (66%) |  |
| West | 12 (11%) |  |
| PAH Etiology, n (%) |  |  |
| Idiopathic | 67 (61%) |  |
| Connective Tissue Disease | 17 (16%) |  |
| Familial | 9 (8%) |  |
| Congenital Heart Disease | 6 (6%) |  |
| Other | 10 (9%) |  |
| WHO Functional Class, n (%) |  |  |
| Class I | 21 (19%) |  |
| Class II | 55 (50%) |  |
| Class III | 17 (16%) |  |
| Unspecified | 16 (15%) |  |
|  | Spearman's rank correlation (r) | P value |
| Correlation of Total Minutes Asleep with: |  |  |
| 6 Minute Walk Distance | 0.06 | 0.589 |
| Daily Steps | -0.16 | 0.1 |
| Sedentary Minutes | **-0.46** | **<0.001** |
| Lightly Active Minutes | -0.17 | 0.076 |
| Fairly Active Minutes | -0.05 | 0.621 |
| Very Active Minutes | 0.02 | 0.570 |
| Correlation of % Light Sleep with: |  |  |
| 6 Minute Walk Distance | -0.24 | 0.065 |
| Daily Steps | -0.19 | 0.076 |
| Sedentary Minutes | **0.22** | **0.042** |
| Lightly Active Minutes | **-0.32** | **0.003** |
| Fairly Active Minutes | -0.15 | 0.172 |
| Very Active Minutes | -0.02 | 0.836 |
| Correlation of % Deep Sleep with: |  |  |
| 6 Minute Walk Distance | **0.32** | **0.012** |
| Daily Steps | **0.30** | **0.005** |
| Sedentary Minutes | **-0.38** | **<0.001** |
| Lightly Active Minutes | **0.42** | **<0.001** |
| Fairly Active Minutes | **0.27** | **0.011** |
| Very Active Minutes | 0.19 | 0.073 |
| Correlation of % REM Sleep with: |  |  |
| 6 Minute Walk Distance | **0.27** | **0.032** |
| Daily Steps | **0.26** | **0.016** |
| Sedentary Minutes | **-0.26** | **0.013** |
| Lightly Active Minutes | **0.32** | **0.003** |
| Fairly Active Minutes | **0.23** | **0.03** |
| Very Active Minutes | 0.27 | 0.054 |

Data are median (IQR) for continuous variables and frequency (percentage) for categorical variables. Fitbit activity and sleep metrics were measured daily for approximately 12 weeks and averaged over the entire monitoring period. Spearman's rank correlation test was used to evaluate associations between sleep and activity metrics in the patients who completed the baseline monitoring period (n=109).
Abbreviations: PAH = pulmonary arterial hypertension. WHO = World Health Organization

### Activity and Sleep Levels

In the PAH cohort who completed the 12-week baseline monitoring period (n=109), the average daily steps were 5202 (IQR 3301 – 6771). Average sedentary minutes per day were 799 (IQR 708-904), lightly active minutes 202 (161 – 260), fairly active minutes 4.8 (1.8 – 8.9) and very active minutes 2.1 (0.4 – 6.6). Average total minutes asleep per day were 399 (IQR 342 – 434). The percentage of time spent in light sleep was 62.7% (58.5 – 66.0), deep sleep 15.0% (12.3 – 17.1), and REM sleep 18.3% (15.3 – 21.6).

### Activity and 6MWD Correlations

There was a significant association between the 6MWD and all Fitbit-derived activity metrics. 6MWD correlated with average daily steps (r = 0.66, p< 0.001, **Figure 1)**, daily sedentary minutes (r= -0.37, p < 0.001), daily lightly active minutes (r = 0.51, p = < 0.001), daily fairly active minutes (r = 0.44, p < 0.001) and daily very active minutes (r=0.56, p < 0.001).

**Figure 1:**
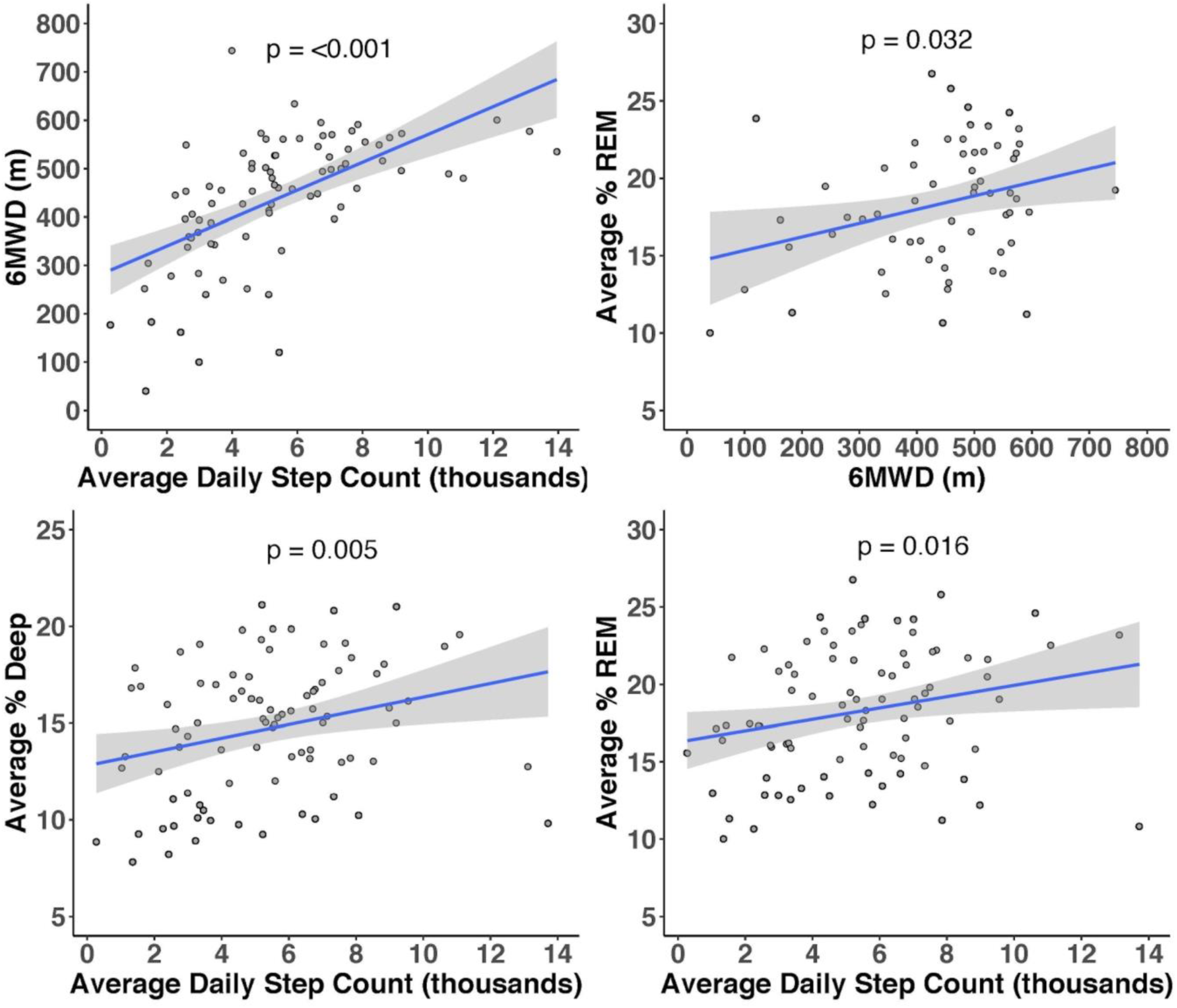
Correlation Plots for Baseline Sleep and Activity Measures. In the cohort of patients with PAH who completed a 12-week baseline monitoring period (n=109), correlation plots demonstrated significant associations between several sleep and activity metrics.

### Activity and Sleep Correlations

We observed a significant positive correlation between percent of time spent in restorative sleep stages (% Deep and % REM sleep) and average daily steps, 6MWD, lightly active minutes and fairly active minutes as well as an inverse relationship between these high-quality sleep stages and sedentary minutes **(Table 1**, **Figure 1).** Total minutes asleep was only associated with sedentary minutes (r = -0.46, p<0.001).

### Activity and QOL Correlations

Average daily steps showed a significant negative correlation with MLHF total score (r = -0.4, p = 0.010) and MLHF physical score (r = -0.42, p = 0.006). Since higher MLHF scores indicate greater impairment, this shows that steps are lower in patients with worse QOL. The correlation between average daily steps and MLHF emotional score was r = -0.31 (p = 0.052). Sedentary minutes correlated with the MLHF total score (r = 0.44, p = 0.004) and MLHF physical score (r=0.36, p = 0.019) but was not associated with MLHF emotional score (r=0.24, p=0.133). Lightly active minutes correlated with MLHF emotional score (r = -0.33, p = 0.040) but did not correlate with MLHF total score (r = -0.25, p= 0.118) or MLHF physical score (r = -0.22, p=0.153). Fairly active minutes were negatively correlated with all three MLHF scores. Very active minutes were associated with MLHF total score (r = -0.42, p = 0.007) and MLHF physical score (r = -0.41, p = 0.007), but not MLHF emotional score (r=-0.21, p=0.187).

### Sleep and QOL Correlations

There were no significant associations between sleep metrics and MLHF QOL scores.

### Longitudinal Trends in PAH Patients

44 out of the 109 participants completed both a 12-week monitoring period at baseline and one-year follow-up. The median time between the start of the baseline monitoring period and the follow-up monitoring period was 1.03 years (IQR 0.96 – 1.34 years). The cohort composition was similar with a median age of 55.2 years old (43.3 – 62.1), 80% female, 59% WHO functional class II, and 59% idiopathic PAH **(Table 2)**

**Table 2:**
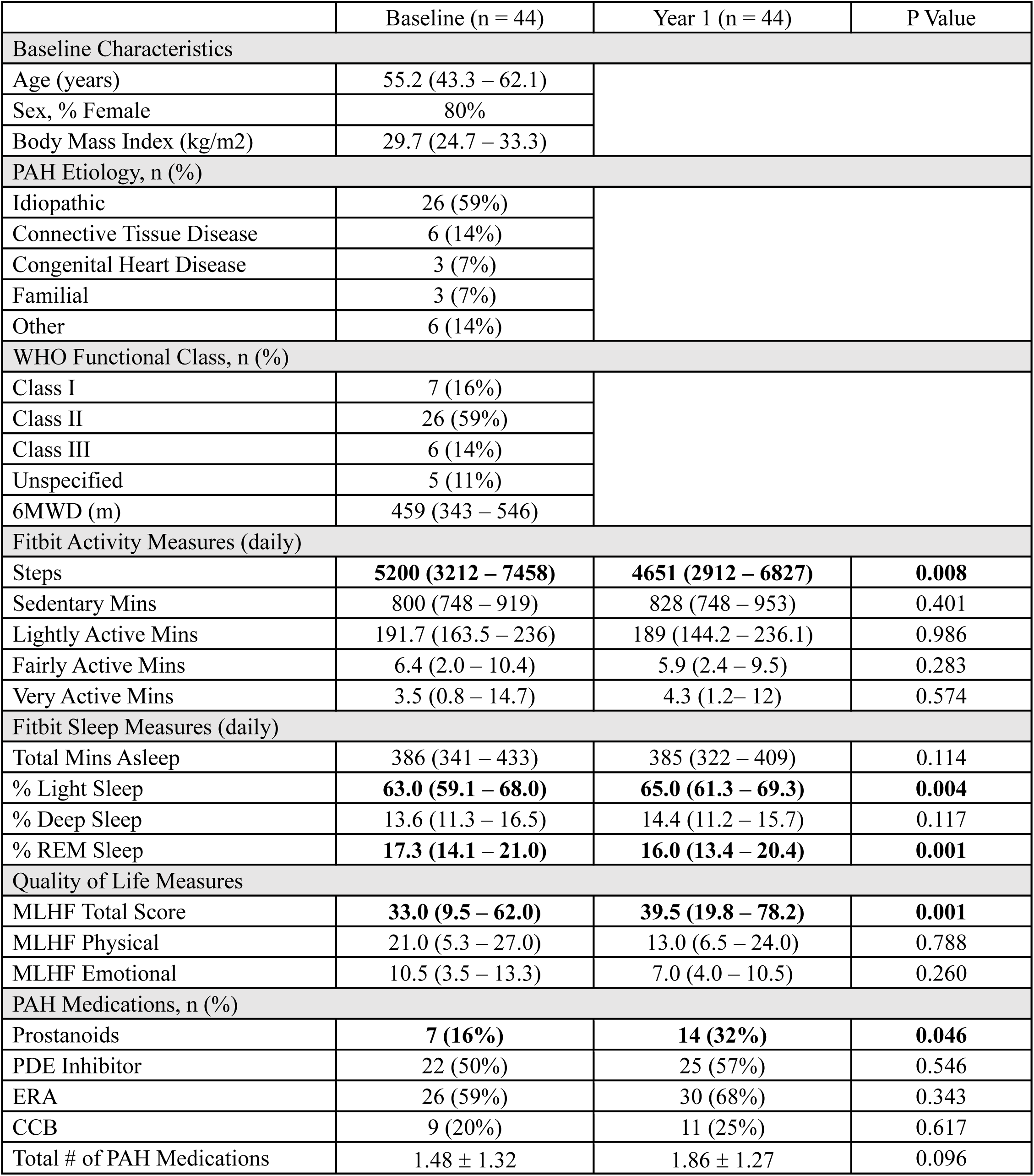
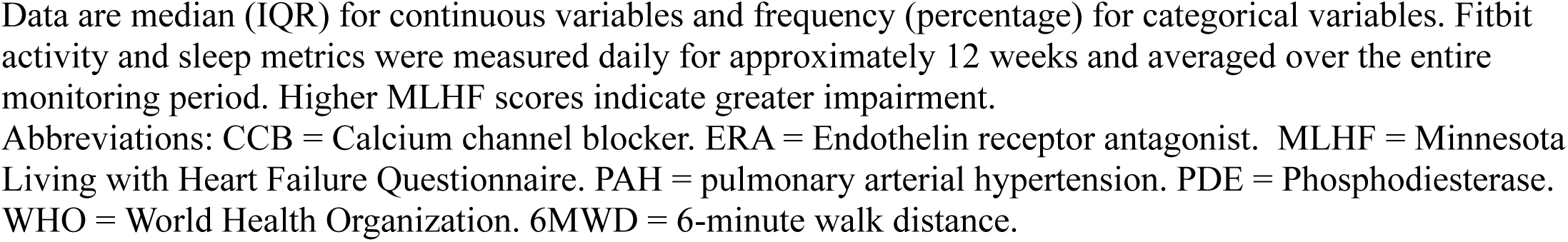
Changes in Activity, Sleep, Quality of Life, and Medications in Patients with PAH.

|  | Baseline (n = 44) | Year 1 (n = 44) | P Value |
| --- | --- | --- | --- |
| Baseline Characteristics |  |  |  |
| Age (years) | 55.2 (43.3 – 62.1) |  |  |
| Sex, % Female | 80% |  |  |
| Body Mass Index (kg/m2) | 29.7 (24.7 – 33.3) |  |  |
| PAH Etiology, n (%) |  |  |  |
| Idiopathic | 26 (59%) |  |  |
| Connective Tissue Disease | 6 (14%) |  |  |
| Congenital Heart Disease | 3 (7%) |  |  |
| Familial | 3 (7%) |  |  |
| Other | 6 (14%) |  |  |
| WHO Functional Class, n (%) |  |  |  |
| Class I | 7 (16%) |  |  |
| Class II | 26 (59%) |  |  |
| Class III | 6 (14%) |  |  |
| Unspecified | 5 (11%) |  |  |
| 6MWD (m) | 459 (343 – 546) |  |  |
| Fitbit Activity Measures (daily) |  |  |  |
| Steps | 5200 (3212 – 7458) | 4651 (2912 – 6827) | 0.008 |
| Sedentary Mins | 800 (748 – 919) | 828 (748 – 953) | 0.401 |
| Lightly Active Mins | 191.7 (163.5 – 236) | 189 (144.2 – 236.1) | 0.986 |
| Fairly Active Mins | 6.4 (2.0 – 10.4) | 5.9 (2.4 – 9.5) | 0.283 |
| Very Active Mins | 3.5 (0.8 – 14.7) | 4.3 (1.2– 12) | 0.574 |
| Fitbit Sleep Measures (daily) |  |  |  |
| Total Mins Asleep | 386 (341 – 433) | 385 (322 – 409) | 0.114 |
| % Light Sleep | 63.0 (59.1 – 68.0) | 65.0 (61.3 – 69.3) | 0.004 |
| % Deep Sleep | 13.6 (11.3 – 16.5) | 14.4 (11.2 – 15.7) | 0.117 |
| % REM Sleep | 17.3 (14.1 – 21.0) | 16.0 (13.4 – 20.4) | 0.001 |
| Quality of Life Measures |  |  |  |
| MLHF Total Score | 33.0 (9.5 – 62.0) | 39.5 (19.8 – 78.2) | 0.001 |
| MLHF Physical | 21.0 (5.3 – 27.0) | 13.0 (6.5 – 24.0) | 0.788 |
| MLHF Emotional | 10.5 (3.5 – 13.3) | 7.0 (4.0 – 10.5) | 0.260 |
| PAH Medications, n (%) |  |  |  |
| Prostanoids | 7 (16%) | 14 (32%) | 0.046 |
| PDE Inhibitor | 22 (50%) | 25 (57%) | 0.546 |
| ERA | 26 (59%) | 30 (68%) | 0.343 |
| CCB | 9 (20%) | 11 (25%) | 0.617 |
| Total # of PAH Medications | 1.48 ± 1.32 | 1.86 ± 1.27 | 0.096 |

Data are median (IQR) for continuous variables and frequency (percentage) for categorical variables. Fitbit activity and sleep metrics were measured daily for approximately 12 weeks and averaged over the entire monitoring period. Higher MLHF scores indicate greater impairment.
Abbreviations: CCB = Calcium channel blocker. ERA = Endothelin receptor antagonist. MLHF = Minnesota Living with Heart Failure Questionnaire. PAH = pulmonary arterial hypertension. PDE = Phosphodiesterase. WHO = World Health Organization. 6MWD = 6-minute walk distance.

Average daily steps decreased from 5200 [IQR 3212 – 7458] at baseline to 4651 [IQR 2912 – 6827] at one year, (p = 0.008), but the active minutes did not change significantly (**Table 2).** The percent of time in light sleep increased from 63.0% [59.1% - 68.0%] at baseline to 65.0% [61.3 – 69.3%] at one year, (p = 0.004), and the percent of time in REM sleep decreased from 17.3% [14.1 – 21.0] to 16.0% [13.4 – 20.4] (p = 0.001) (**Table 2**, **Figure 2).** MLHF Total Score increased significantly from 33.0 (IQR 9.5 – 62.0) at baseline to 39.5 (19.8 – 78.2) at one year (p=0.001), which illustrates worse QOL at one year. MLHF physical and MLHF emotional scores were not significantly changed over this time. The number of participants on prostanoids increased from 16% at baseline to 32% at one year follow-up, (p = 0.046), but there was no significant change in the total number of PAH medications per participant at one-year.

**Figure 2:**
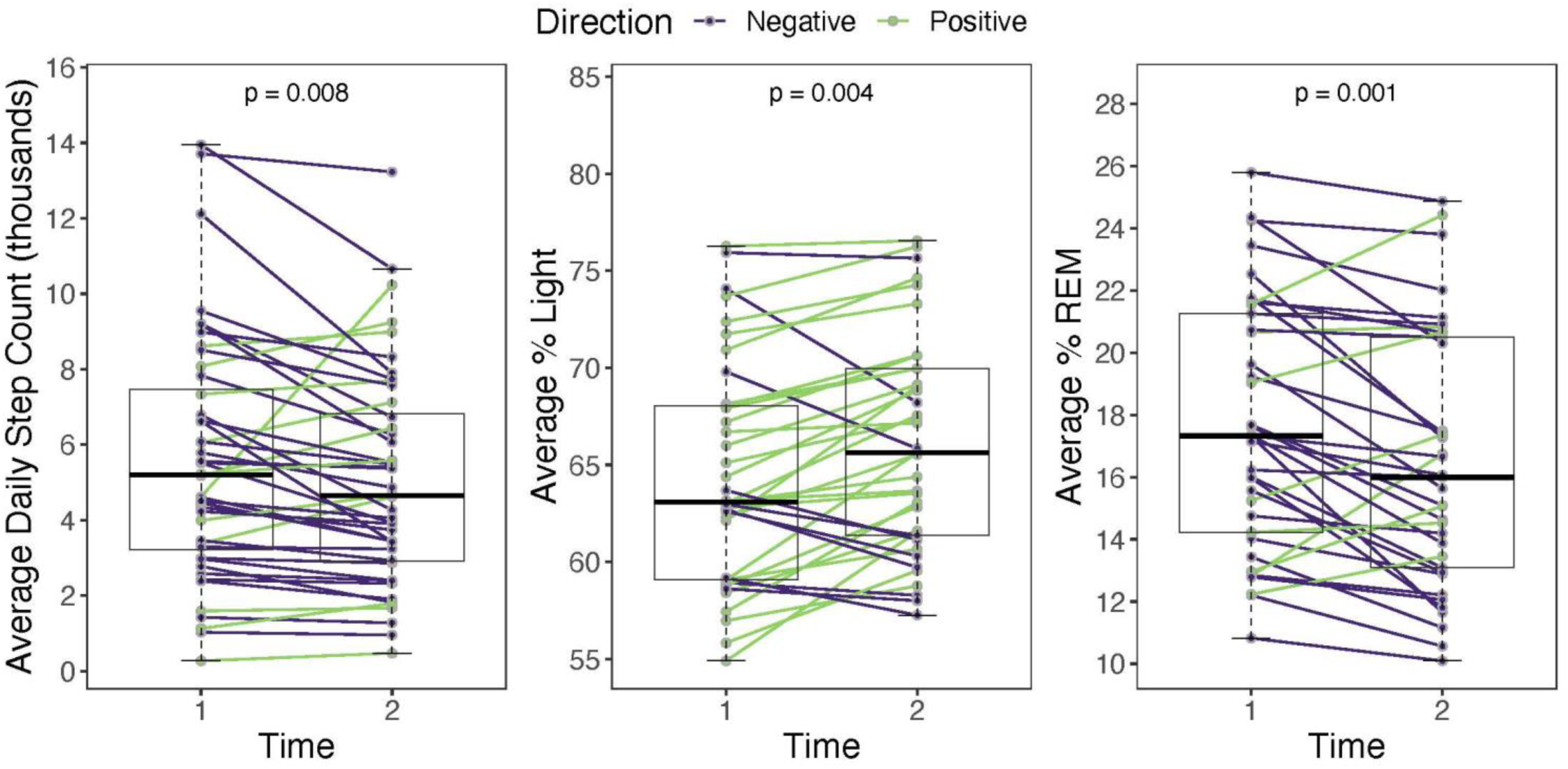
Longitudinal Activity and Sleep Metrics. In the PAH cohort with baseline and one-year follow-up (n=44), spaghetti plots demonstrate an overall decrease in average daily steps, an increase in the percent light sleep per night, and a decrease in the percent REM sleep per night. Green lines denote individuals with an increased value from baseline to year 1 while purple lines denote individuals with a decreased value from baseline to year 1.

### Activity Levels in PAH and Matched Controls

Average daily steps and active minutes were significantly lower in patients with PAH than matched controls at baseline and one-year follow-up. Average sedentary minutes were significantly higher in patients with PAH compared to matched controls at both time points (Table 3).

**Table 3:** Activity Levels and Sleep Metrics in Patients with PAH and Matched Controls.

|  | PAH Cohort<br>(n = 44) | Matched Controls<br>(n = 132) | P Value |
| --- | --- | --- | --- |
| **Baseline Characteristics** |  |  |  |
| Age (years) | 55.2 (43.3 – 62.1) | 53.8 (41.4 – 61.9) | 0.502 |
| Sex, % Female | 80% | 80% | 1 |
| Body Mass Index (kg/m<sup>2</sup>) | 29.7 (24.7 – 33.3) | 29.3 (24.2 – 33.8) | 1 |
| **Activity Measurements** |  |  |  |
| Daily Steps: Baseline | **5200 (3212 – 7458)** | **7369 (5225 – 10089)** | **< 0.001** |
| Daily Steps: Year 1 | **4651 (2912 – 6827)** | **7044 (5010 – 9834)** | **< 0.001** |
| Sedentary Mins: Baseline | **800 (748 – 919)** | **674 (623 – 745)** | **< 0.001** |
| Sedentary Mins: Year 1 | **828 (748 – 953)** | **690 (630 – 734)** | **< 0.001** |
| Lightly Active Mins: Baseline | **191.7 (163.5 – 236.0)** | **219.6 (182.9 – 268.4)** | **0.009** |
| Lightly Active Mins: Year 1 | **189.0 (144.2 – 236.1)** | **221.1 (186.8 – 255.0)** | **0.004** |
| Fairly Active Mins: Baseline | **6.4 (2.0 – 10.4)** | **14.0 (7.3 – 23.7)** | **< 0.001** |
| Fairly Active Mins: Year 1 | **5.9 (2.4 – 9.5)** | **13.5 (6.9 – 22.2)** | **< 0.001** |
| Very Active Mins: Baseline | **3.5 (0.8 – 14.7)** | **16.3 (6.0 – 28.5)** | **< 0.001** |
| Very Active Mins: Year 1 | **4.3 (1.2 – 12.1)** | **13.4 (5.7 – 25.1)** | **< 0.001** |
| **Sleep Measurements** |  |  |  |
| Mins Asleep: Baseline | 386 (341 – 433) | 395 (367 – 424) | 0.518 |
| Mins Asleep: Year 1 | **385 (322 – 409)** | **397 (370 – 425)** | **0.039** |
| % Light Sleep: Baseline | 63.0 (59.1 – 68.0) | 63.3 (59.8 – 68.1) | 0.921 |
| % Light Sleep: Year 1 | 65.0 (61.3 – 69.3) | 63.7 (60.2 – 67.9) | 0.194 |
| % Deep Sleep: Baseline | 13.6 (11.3 – 16.5) | 15.2 (12.8 – 17.5) | 0.095 |
| % Deep Sleep: Year 1 | **14.4 (11.2 – 15.7)** | **15.2 (13.1 – 17.3)** | **0.02** |
| % REM Sleep: Baseline | **17.3 (14.1 – 21.0)** | **21.6 (18.7 – 23.8)** | **< 0.001** |
| % REM Sleep: Year 1 | **16.0 (13.4 – 20.4)** | **21.3 (17.9 – 23.8)** | **< 0.001** |

Data are median (IQR) for continuous variables and frequency (percentage) for categorical variables. Fitbit activity and sleep metrics were measured daily for approximately 12 weeks and averaged over the entire monitoring period.

### Sleep Patterns in PAH and Matched Controls

Total minutes asleep were similar in patients with PAH and matched controls at baseline. Patients with PAH had shorter average sleep duration at one year compared to controls (385 minutes (322 – 409) vs 397 minutes (370 – 425), p = 0.039; **Table 3**). In the adjusted models, patients with PAH had, on average, 36 (14 – 57) fewer minutes of sleep at one-year compared to controls (p=0.001; **Figure 3**). Percent light sleep was similar in patients with PAH and matched controls at baseline and one year follow-up. In adjusted models, there was a significant increase in % light sleep in patients with PAH from baseline to year 1 (p=0.004), but no significant change in controls (p = 0.534). We also observed a group-time interaction (p = 0.024), which shows that % light sleep increased significantly over the course of one year in patients with PAH compared to controls. Average % REM sleep was significantly lower in patients with PAH compared to controls at baseline and year 1. The adjusted models showed that % REM decreased significantly in patients with PAH from baseline to year 1 (p=0.002), it did not decline significantly in controls, and there was a significant group-time interaction confirming that relationship over one year (p =0.034).

**Figure 3:**
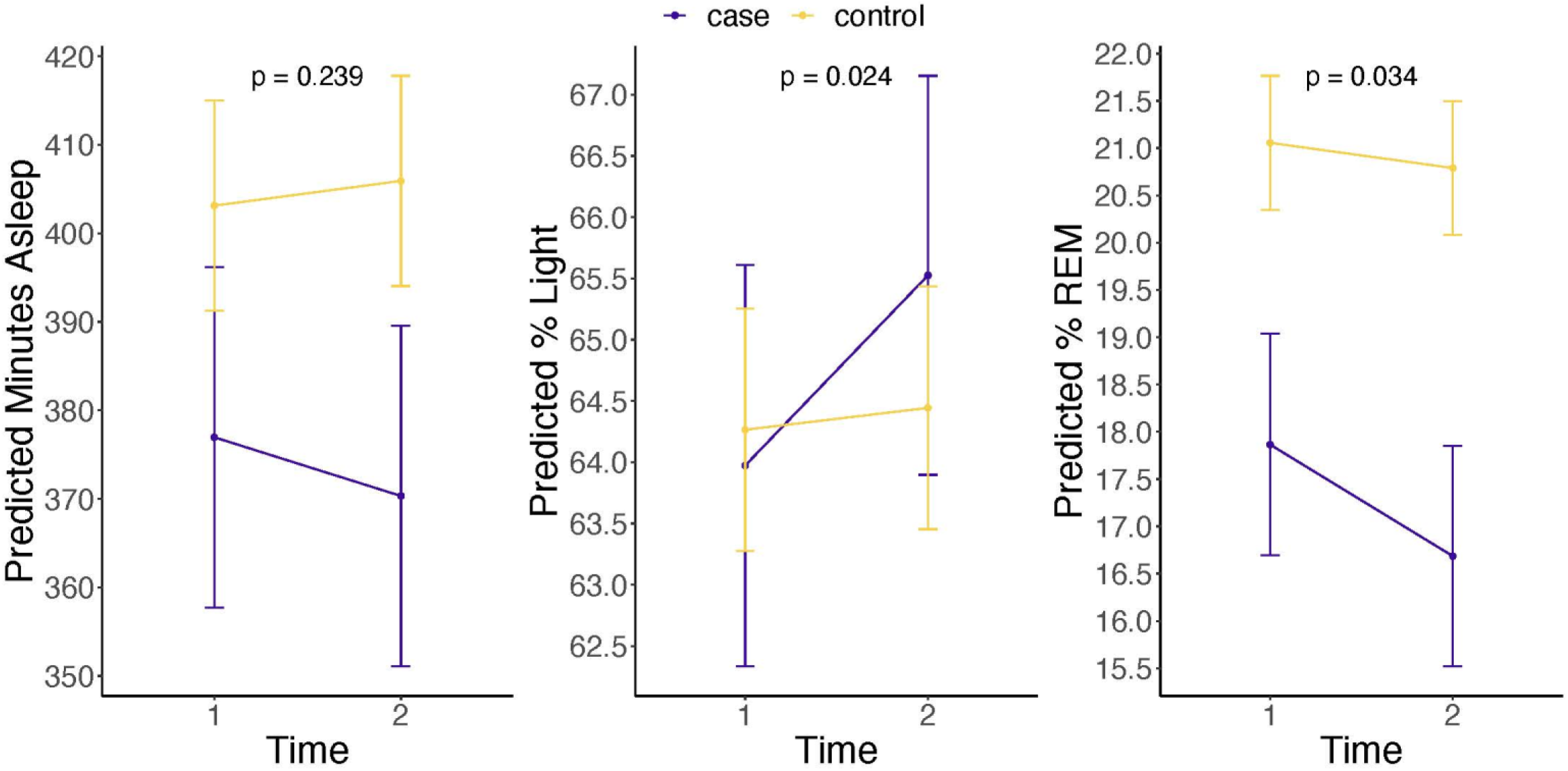
Comparison of Sleep Duration and Sleep Quality in Patients with PAH and Matched Controls. Generalized least squares models, adjusted for age, sex, and body mass index (BMI), with linear interaction analyses evaluated changes in total minutes asleep, percent light sleep, and percent REM sleep over the course of one year. The group-time interaction term was significant for percent light sleep (p = 0.024) and percent REM sleep (p = 0.034), which indicates that sleep quality worsened in patients with PAH over time compared to matched controls. The interaction was not significant for total minutes asleep (p = 0.239).

## Discussion

In this study, we demonstrate that daily activity and sleep quality are correlated, activity levels and sleep quality are significantly worse in patients with PAH compared to matched controls, and these measures decline over one year in patients with PAH. These reflect important health behaviors that are not captured in routine clinic-based measurements. Our study provides novel insights into sleep quality as it is the first to report longitudinal sleep-stage patterns in PAH and the first to investigate sleep and activity in the context of rigorously matched controls.

Functional capacity is a strong prognostic marker in PAH^16,17^. 6MWD is the most common measure of functional capacity but it has important limitations as a clinical surveillance measure^3^ with a narrow range of values, ceiling and floor effects^18,19^, and is vulnerable to transient influences at the moment of testing. Free-living activity monitoring may overcome these limitations. We found that all Fitbit-derived activity metrics correlated with the 6MWD, but the R^2^ was between 0.1 and 0.35. This shows that, at best, the 6MWD accounts for a third of the variability in daily activity. 6MWD and wearable devices provide different, complementary information as real-world activity monitoring incorporates behavioral choices as well as social and environmental influences. Additionally, our study found that daily steps declined over one year, but sedentary and active minutes did not change. Daily steps in PAH appear to represent modifiable health behaviors as a text-based mobile health intervention significantly increased steps and QOL in PAH without improving 6MWD^12^. This supports the notion that steps are a clinically meaningful measure that captures complementary information to clinic-based measurements, but future adequately powered studies are needed to investigate whether declining steps precede clinically significant outcomes.

To our knowledge, this is the first study to evaluate long-term sleep patterns in patients with PAH using wearable devices that capture sleep stages. Prior work suggests sleep is important in PAH but existing studies were limited by self-report qualitative surveys or single night sleep studies^6,7,20^. Sleep health is an important and underrecognized component of cardiovascular health^21^ with emerging data that wearable-based sleep data (duration, irregularity, and quality) are important factors that may contribute to the development of many chronic conditions^22^. In the general population, decreased REM sleep has been associated with a higher risk of all-cause and cardiovascular mortality^23^, and more REM sleep may be protective against incident heart failure^24^. Sleep health warrants increased attention in PAH, especially given the high prevalence of sleep disordered breathing^25^ and high self-reported rates of poor sleep quality^6^. We found that sleep quality and activity measures were modestly but consistently correlated. We observed that sleep quality in patients with PAH worsened over time compared to matched controls as evident by an increased percentage of time in light sleep and decreased percentage of time in REM sleep. The decreased sleep quality that we observed in patients with PAH may arise from physical symptoms while awake, mental health symptoms which are highly prevalent in PAH, or sleep disordered breathing conditions. Nocturnal desaturations are highly prevalent and underrecognized in this population and may be a modifiable contributor to poor sleep quality^20^.

The main strength of our study is its ability to use objective “real-world” data from wearable devices to describe health behaviors over time in a large cohort of patients with PAH compared to matched controls. While physical activity and sleep have historically been examined separately, there is emerging evidence that these behaviors are inter-related and exist along one movement continuum^26,27^. This is the first study to describe the inter-related nature of sleep and activity behaviors in PAH. The association between physical activity, sedentary behavior, and sleep suggests that clinical interventions to improve one health behavior may positively impact the other health behaviors as well. Additionally, our PAH cohort reported worse QOL over the course of one year. This suggests that the decreased sleep quality and daily steps in our cohort may also capture subclinical changes in QOL, which may not be reflected in traditional clinic-based assessments. Future studies are warranted to determine whether real-time, continuous monitoring of physical activity and sleep can detect a “digital prodrome” of functional decline that could improve outcomes by triggering medical contact. This is conceptually similar to the use of CardioMEMS devices to detect and act on increases in filling pressure^28–30^. Our study highlights the potential value of integrating these health behaviors into future studies, such as n-of-1 trials targeting specific adverse behaviors (i.e. poor sleep quality/duration or reduced activity).

The findings of our study should be viewed in the context of several limitations. We used commercially available Fitbit devices which have reduced fidelity compared with research-grade actigraphy, but the use of commercial devices increases generalizability as many patients already use these devices in their daily lives. Our PAH cohort was quite functional at baseline as 70% were functional class I-II and they were only on an average of 1.5 PAH medications, so their activity and sleep quality may decline at a different rate than PAH patients who were later in their disease course. Nevertheless, we were still able to show that step count and sleep quality decline over one year despite enrolling patients with more mild disease. Another limitation is that only 40% of participants completed the one-year follow-up, which occurred for multiple reasons. Twenty-two participants were included in the baseline analyses from a trial that by design did not have a follow-up period. Additionally, the study is ongoing so many participants were recently enrolled and may complete their one-year follow-up at a later date. Given the geographic diversity of the PAH cohort, it is possible that there is some seasonal and geographic variation that we were unable to account for with the matched controls. Lastly, some data was collected during the COVID-19 pandemic which may have decreased activity levels and changed sleep patterns. The matched monitoring dates in the control subjects mitigates the impact of the pandemic, but patients with chronic cardiopulmonary conditions may have been more adherent to social isolation guidelines compared to the controls thereby decreasing their activity levels.

In conclusion, this study illustrates the potential clinical value of activity and sleep monitoring with wearable devices and provides novel insights pertaining to health behaviors in patients with PAH. Activity and sleep quality are reduced in patients with PAH compared with matched controls and decline over one year. Sleep quality and physical activity may represent modifiable and novel targets that could improve how patients with PAH feel and function. Future studies are warranted to test whether interventions to improve sleep and activity lead to better outcomes and whether real-time continuous monitoring of sleep and activity behaviors can detect early functional decline.

## Data Availability

Data for this study can be made available through data use agreements with Vanderbilt and the PVDOMICS program. All data from All of Us are publicly available.

## Abbreviations

AoURP: All of Us Research Program
BMI: Body Mass Index
L-PVDOMICS: Longitudinal Pulmonary Vascular Disease Phenomics Program
MLHF: Minnesota Living with Heart Failure Questionnaire
PAH: Pulmonary Arterial Hypertension
QOL: Quality of Life
REM: Rapid Eye Movement
WHO: World Health Organization
6MWD: 6 Minute Walk Distance
6MWT: 6 Minute Walk Test

## Acknowledgements

None

